# Morphological feature remodeling of intracranial arteries in the context of inflammation and HIV-associated cognitive impairment

**DOI:** 10.64898/2026.05.19.26353071

**Authors:** Nhat Hoang, Hongmei Yang, Md Nasir Uddin, Jianhui Zhong, Abrar Faiyaz, Meera V. Singh, Zachary D. Boodoo, Karli Sutton, Henry Z. Wang, Bogachan Sahin, Mohammed W. Khan, Miriam T. Weber, Chun Yuan, Li Chen, Giovanni Schifitto

## Abstract

**Background:** Despite the success of combination antiretroviral therapy (cART), vascular comorbidities, including cerebrovascular disease, are more prominent in people living with HIV (PLWH) compared to people without HIV (PWOH). However, quantitative assessments of cerebrovascular morphometry and their associations with cognitive outcomes in the context of HIV are still limited. In this study, we explore this missing link.

**Methods:** Magnetic Resonance Angiography (MRA) data, blood markers, and neurocognitive assessments were collected from 73 PWOH subjects (*male*: 57, *female*: 16; *age*: 53 *±* 16) and 99 PLWH subjects (*male*: 66, *female*: 30, *age*: 53 *±* 11). Vessel morphometric features were quantified using intraCranial Artery Feature Extraction (iCafe) to investigate associations between vessel morphometry, markers of monocytes, endothelial cell activation, and cognitive performance.

**Results:** HIV status predicted a lower total number of branches (*β* = *−*0.224, *p* = 0.001, *d* = *−*0.517) and shorter total distal length (*β* = *−*0.173, *p =* 0.021, *d =* –0.370) with a moderate effect size. Total branch number was found to be negatively associated with plasma levels of monocyte markers (sCD14: *r =* –0.167, *p =* 0.033; sCD163: *r =* –0.157, *p =* 0.045) and positively correlated with white matter cerebral blood flow (*r =* 0.550; *p ≤* 0.05). HIV status was the strongest predictor of overall cognitive performance in ANCOVA model (*β* = –0.219, *p =* 0.006, *d =* –0.453).

**Conclusions:** Our results suggest that cognitive impairment in PLWH is associated with vessel morphology metrics. Monocyte immune activation may contribute to changes in vessel morphology.

## Introduction

Although combination antiretroviral therapy (cART) has successfully prolonged the life expectancy of people living with HIV (PLWH), they continue to face an increased risk of neurological complications due to the persistence of chronic inflammation ^1–3^. Therefore, aging PLWH face the consequences of the combined effect of HIV and aging-mediated inflammation. Accordingly, growing evidence suggests that PLWH experience a greater burden of age-related conditions, such as cardiovascular and cerebrovascular disease ^4–8^.

Quantitative metrics of intracranial vessels provide an opportunity to assess the extent of vascular pathology in PLWH. Three-dimensional time-of-flight (3D-TOF) magnetic resonance angiography (MRA) is a standard noninvasive angiography technique used in clinical practice ^9^. With advances in image processing, extracting cerebral vascular features from 3D-TOF MRA is now possible. Recently, Chen et al. developed a semi-automated approach called IntraCranial Artery Feature Extraction (iCafe), which can effectively evaluate the intensity and morphometry features of the vascular regions in the brain, using 3D-TOF MRA images ^10^. While conventional TOF MRA provides qualitative information, iCafe models TOF images and generates vascular feature maps capable of detecting subtle changes in all vascular regions ^11^. iCafe-derived features, such as branch number, tortuosity, diameter, length, and vessel density, have been shown to correlate with age ^11^, atherosclerosis ^12^, cerebral small vessel disease (CSVD) ^13^, and cerebral blood flow (CBF) ^14^. Tortuosity, length, volume, and vessel branch features generated by similar techniques to iCafe also revealed significant association with acute ischemic stroke and Alzheimer’s Disease (AD) ^15^.

In this study, we used iCafe to measure cerebrovascular burden in PLWH. We hypothesized that iCafe metrics would differ between PLWH and people without HIV (PWOH) and vary with plasma levels of monocyte and endothelial activation. Additionally, we assessed the impact of iCafe metrics on cerebral blood flow and cognitive performance.

## Methods

### 1. Study Participants

Prior to enrollment, all participants provided written informed consent in accordance with the protocols of the Research Subject Review Board (RSRB) at the University of Rochester. Participants underwent clinical, laboratory, brain MRI, and neuropsychological assessment. A detailed description of the study protocol has been previously published ^16^.

### 2. Blood specimen collection

A 40 mL whole-blood sample was collected in sterile Acid Citrate Dextrose (ACD) vacutainers by venipuncture. Blood was processed within 2 hours of collection and was incubated at room temperature on a slow shaker until processing. Plasma was isolated by centrifugation at 1000 x G for 10 minutes, and cryopreserved at -80 degrees Celsius before being processed with enzyme-linked immunosorbent assay (ELISA) for monocyte activation markers, soluble CD14 (sCD14, R and D systems, catalog DY383-05) and CD163 (R and D systems, catalog DY1607-05) as well as endothelial activation markers intracellular adhesion molecule 1 (ICAM, R and D systems, catalog DY720-05), osteoprotegerin (R and D systems, catalog DY805) and vascular adhesion molecule (VCAM, R and D systems, catalog DY809-05) as described in manufacturer’s protocol.

### 3. Neurocognitive assessment

All participants were assessed on a battery of neuropsychological tests that included the following cognitive domains: executive function (Trailmaking Test Parts A & B, Stroop Interference task), speed of information processing (Symbol Digit Modalities Test and Stroop Color Naming), attention and working memory (CalCAP CRT4), learning (Rey Auditory Verbal Learning Test Total Learning, Rey Complex Figure Test Immediate Recall), memory (Rey Auditory Verbal Learning Test Delayed Recall, Rey Complex Figure Test Delayed Recall), language (Controlled Oral Word Association Test), and motor function (Grooved Pegboard). Neurocognitive tests were administered by staff trained and supervised by a licensed clinical neuropsychologist. Raw scores were converted to z-scores using standardized norms corrected for age and education. Domain-specific cognitive scores were then calculated by averaging the z-scores of tests within each domain. Finally, a total cognitive z-score was computed by summing all the domain-specific z-scores to provide an overall assessment of cognitive performance.

### 4. MRA and MRI acquisition

All MR images were collected on a 3T Siemens (Erlangen, Germany) MAGNETOM PrismaFit whole-body scanner equipped with a 64-channel phased array coil. T1-weighted (T1w) anatomical images were acquired with a three-dimensional magnetization prepared rapid acquisition gradient-echo (3D MPRAGE) sequence with inversion time (TI) = 926 ms, repetition time (TR) = 1840 ms, echo time (TE) = 2.45 ms, flip angle = 8°, field of view (FOV) = 256 × 256, GRAPPA = 2, image resolution = 1.0 × 1.0 × 1.0 mm^3^. 3D fluid attenuated inversion recovery (3D FLAIR) images were collected with TI=1800 ms, T*R =* 5000 ms TE= 215 ms, image resolution = 1.0 × 1.0 × 1.0 mm^3^. MRA images were acquired with a 3D time of flight (TOF) MRA sequence using T*R =* 21 ms, TE= 3.42 ms, flip angle = 20°, GRAPPA=2, phase oversampling=30%, slice oversampling=20.0 %, image resolution =0.5 × 0.5 × 0.5 mm^3^. To quantify cerebral blood flow (CBF), 2D multiband pseudo continuous arterial spin labeling (ASL) was acquired using a 2D echo planar imaging with multiband acceleration facto*r =* 6, T*R =* 3594 ms, TE = 19 ms, flip angle = 90°, FOV = 215 × 215, image resolution = 2.5 × 2.5 × 2.3 mm^3^.

### 5. Data processing

#### 5.1 iCafe artery tracing and feature extraction

We employed the iCafe tool^17^ to extract vascular features. iCafe utilizes a center-line tracing algorithm to model and compute vascular information for 22 arterial regions within the circle of Willis (CoW), the basilar arteries (BA), and the vertebral arteries (VA) ^10^. The full notation list of vascular segment abbreviations is included in the Footnote/Abbreviation section.

After the vessel tracings are generated, a trained operator then removes any false tracings and marks regions of interest to prepare for semi-automatic vessel labeling. This procedure for each subject is quality-controlled by two vascular neurologists and a neuroradiologist to minimize the presence of faulty vessel branches and mislabeling during segmentation. Additionally, cross-validation between quality-controlled subjects was performed to ensure reproducibility of the results.

Figure 1 illustrates the iCafe artery tracing procedure for two age-matched subjects in age group 21-25, one from the PLWH cohort and the other from the PWOH. Post-segmentation, iCafe calculates vessel features using a maximum a posteriori estimation and transfers the processed data to a SQL server for feature extraction.

**Figure 1:**
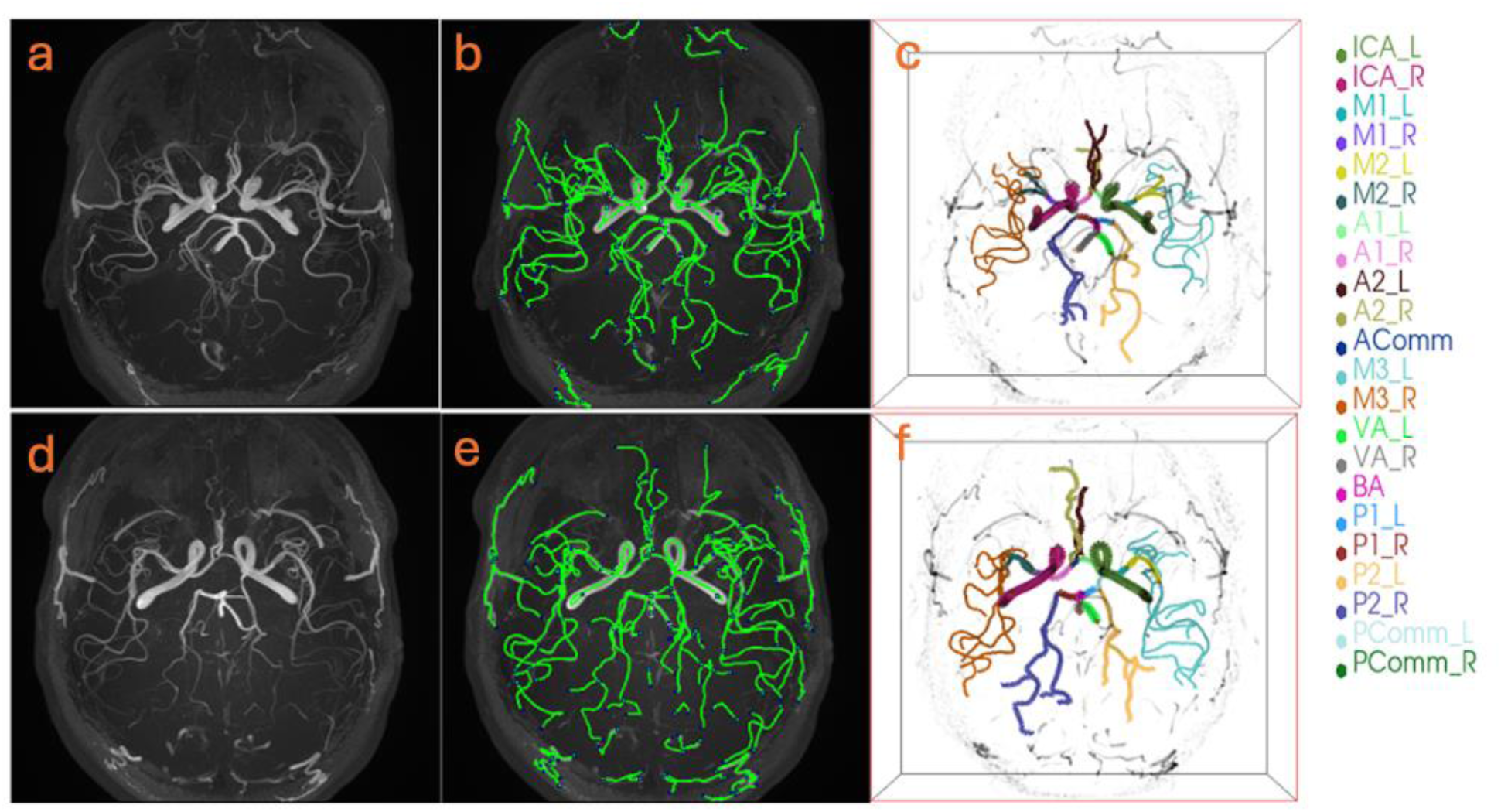
iCafe pipeline comparison between a pair of age-matched HIV- and HIV+ subjects. The iCafe tracing between two age-matched subjects in age group 21-25, with the PWOH subject on the top row and the PLWH subject on the bottom row. a) and d): Maximum intensity projection of MRA scans. b) and e): iCafe vascular skeleton, c) and f): Final iCafe artery tracing result indicating different intracranial arteries with assorted colors. A list of artery abbreviations is provided in the footnote.

#### 5.2 iCafe feature selection

We focused on vessel branch number, length, diameter, and tortuosity as the primary iCafe vascular features. Branch number refers to the number of branches detected by iCafe for a labeled artery/artery of interest. The total branch number is the sum of all branch numbers in the CoW, plus those in the vertebral arteries and the basilar artery. The average diameter of a labeled artery is calculated as the mean diameter measured along the centerline of each associated branch. The length of the branch segment is defined as the distance along the vessel centerline tracing between the segment end points. The total distal length metric is computed by summing the lengths of all the segments from the MCA, PCA, and ACA branches together. The output of this length metric is reported in the number of voxels the segment occupies, then converted to unit of *mm*. The tortuosity of a branch is defined as the ratio between the segment length along the vessel tracing and the Euclidean distance between the two terminal points of an individual artery segment. The average tortuosity of a labeled artery is the mean tortuosity of all the associated branches. In our study, we chose the *total branch number* to represent branch features, and the *total distal length* to represent length features.

#### 5.3 CBF calculation using ASL

We separated ASL volumes into signal volumes and reference volumes for calibration (label-control pair). Then, the ASL data was processed using Oxford ASL toolbox^18^ to measure the mean CBF in regions of interest, with estimated bolus duration = 1.5s, bolus arrival time = 0.7s, and multiband acquisition facto*r =* 6. The CBF measurements were reported in units of ml/100g/min. Global CBF was defined as the average of all mean CBF measurements in cortical gray matter (CGM), subcortical gray matter (SGM), and generalized white matter (GWM).

#### 5.4 CSVD assessment and WMH volumetrics

The team neuroradiologist evaluated the presence of white matter hyperintensity (WMH) using the Fazekas scoring system^19^. Cerebral small vessel disease (CSVD) status was defined as any Fazekas score of 1 or greater. White matter lesion volumes were quantified using volBrain’s LesionBrain, an online segmentation toolbox that processes T1w and FLAIR input images and provides volumetric information of white matter brain lesions^20^.

### 6. Statistical analysis

We employed statistical analyses to address two primary aims: (1) to evaluate vascular morphometric differences in relation to HIV status and inflammatory markers, and (2) to investigate the biological and functional implications of these changes through perfusion and cognition metrics.

ANCOVA models were employed to adjust for potential cofounders such as age, sex, Reynolds cardiovascular risk scores, and CSVD status (assessed via Fazekas scores) to quantify the HIV impact on vessel morphometry. In analyses restricted to the CSVD-positive subgroup, absolute white matter lesion volume was substituted for Fazekas scores as a covariate. For marginal analysis, iCafe features were compared between PLWH and PWOH using Welch’s t-test.

Associations between inflammatory marker levels and iCafe features were examined using Pearson correlation analysis to assess the role of inflammation in vascular alteration, as well as ANCOVA models with the same covariates described above.

Associations between CBF, HIV and cognitive performance were analyzed using ANCOVA models with sex, Reynolds cardiovascular risk scores, CSVD status, and iCAFE total branch number and total distal length metrics as covariates. Age was not included because cognitive scores were age-standardized. Additionally, Pearson correlation analyses were performed to correlate total branch number and total distal length with regional CBF, age, and cognitive scores for PLWH and PWOH groups. The difference between the fitted PLWH and PWOH slopes was calculated, and its significance is denoted by the interaction p-value.

Multiple test adjustments were not applied for this exploratory study. However, we have included the effect size to quantify the magnitude of observed associations and group differences using Cohen’s *d* calculation. For linear model and ANOVAs, the precise model for Cohen’s *d* for a two-group design is given as 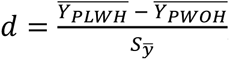, where 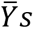 are the adjusted means, and 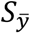 are the adjusted standard deviation. Effect sizes were interpreted using conventional benchmarks (small ≈ 0.2, medium ≈ 0.5, large ≥ 0.8).

The statistical analyses were performed using SAS 9.4 software (SAS Institute Inc, Cary, NC, USA) and *SciPy.Stats* package in Python 3.12 (Python Software Foundation, 2023). Correlation graphs and box plots were generated using Python’s Matplotlib and Seaborn libraries. All p-values <0.05 were considered significant.

## Results

### 1. Demographics and clinical characteristics

Ninety-nine PLWH (*age* 53 *±* 11 years, *female* 30) and 73 PWOH (*age* 53 *±* 16 years, *female* 14) were enrolled. About 10% of PLWH had diabetes compared to 1% of PWOH. Non-Caucasians had a higher representation in the HIV+ group compared to the HIV-group. An overview of the clinical characteristics is provided in Table 1.

**Table 1:**
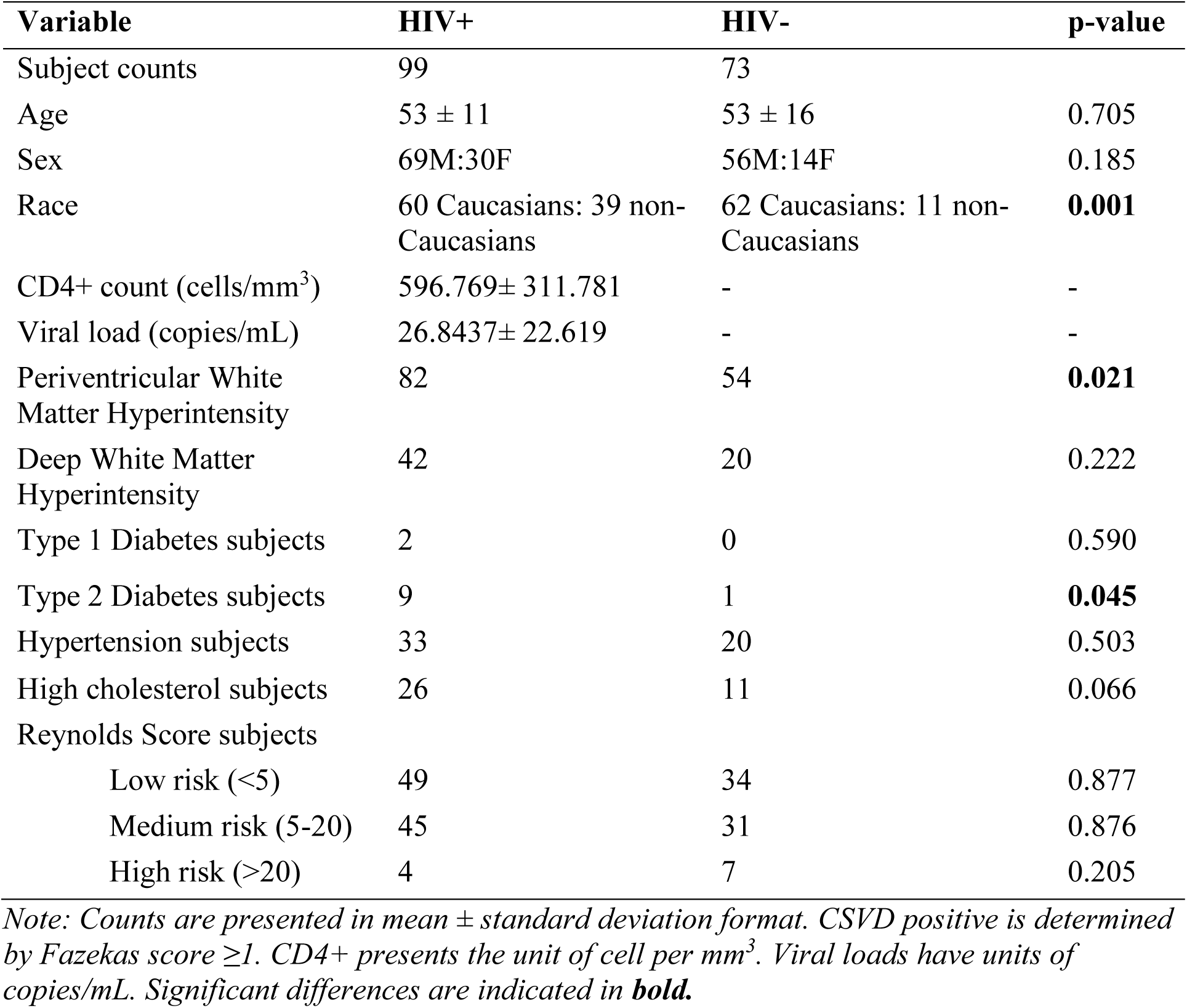
Participants’ demographics and clinical characteristics.

| Variable | HIV+ | HIV- | p-value |
| --- | --- | --- | --- |
| Subject counts | 99 | 73 |  |
| Age | 53 ± 11 | 53 ± 16 | 0.705 |
| Sex | 69M:30F | 56M:14F | 0.185 |
| Race | 60 Caucasians: 39 non-Caucasians | 62 Caucasians: 11 non-Caucasians | **0.001** |
| CD4+ count (cells/mm<sup>3</sup>) | 596.769± 311.781 | - | - |
| Viral load (copies/mL) | 26.8437± 22.619 | - | - |
| Periventricular White Matter Hyperintensity | 82 | 54 | **0.021** |
| Deep White Matter Hyperintensity | 42 | 20 | 0.222 |
| Type 1 Diabetes subjects | 2 | 0 | 0.590 |
| Type 2 Diabetes subjects | 9 | 1 | **0.045** |
| Hypertension subjects | 33 | 20 | 0.503 |
| High cholesterol subjects | 26 | 11 | 0.066 |
| Reynolds Score subjects |  |  |  |
| Low risk (<5) | 49 | 34 | 0.877 |
| Medium risk (5-20) | 45 | 31 | 0.876 |
| High risk (>20) | 4 | 7 | 0.205 |

*Note: Counts are presented in mean ± standard deviation format. CSVD positive is determined by Fazekas score ≥1. CD4+ presents the unit of cell per mm<sup>3</sup>. Viral loads have units of copies/mL. Significant differences are indicated in **bold**.*

### 2. HIV and its interaction with vessel morphometry

#### 2.1. Marginal analysis between PLWH and PWOH

Marginal t-tests revealed that PLWH’s total branch number (58.41 *±* 10.06) was lower compared to the PWOH group (64.23 *±* 14.26; *t = −*2.980, *p =* 0.003, *d = −*0.484). Total distal length demonstrated a similar trend, with PLWH’s total distal length (1426.68 *±* 303.62 *mm*) being significantly shorter than PWOH’s (1555.56 *±* 384.90 *mm*; *t = −*2.369, *p =* 0.019, *d =* **–**0.379). The most pronounced local differences were observed in the MCA regions. MCA branch number for PLWH (32.12 *±* 6.72) was significantly lower than PWOH’s (35.70 *±* 8.66; *t = −*2.938, *p =* 0.004, *d =* –0.471). Similarly, both the left and right MCA length for PLWH (left: 496.13 *±* 122.00 *mm*, right: 483.98 *±* 107.61 *mm)* were significantly shorter than their PWOH’s counterparts (left MCA: 537.05 *±* 121.82 *mm, t = −*2.180, *p =* 0.031, *d = −*0.336; right MCA: 532.36 *±* 136.71 *mm, t = −*2.500, *p =* 0.013, *d = −*0.400).

No other significant differences were found in the means of vessel diameter, volume, and tortuosity between PLWH and PWOH groups (Supplemental Table S1).

#### 2.2. ANCOVA model result

HIV status predicted fewer total number of branches (*β*= *−*0.224, *p =* 0.001, *d =* –0.517) and shorter total distal length (*β* = *−*0.173, *p =* 0.021, *d =* –0.370) with moderate effect size. Age was also independently associated with both total branch number (*β* = *−*0.484, *p <* 0.001, *d = −*0.041) and total distal length (*β* = *−*0.326, *p =* 0.001, *d = −*0.025). CSVD status (as determined by the Fazekas score), sex, and Reynolds score were not significant predictors of vessel morphometry metrics (Table 2).

**Table 2:** Regression model results for associations between HIV status, demographic, and clinical covariates with iCafe total branch number and total distal length metrics.

| Dependent Variable | Predictor | Standardized $\beta$ | p-value | Effect Size d |
| --- | --- | --- | --- | --- |
| **Total Branch Number** | HIV status: Positive vs. Negative | −0.224 | **0.001** | −0.517 |
|  | Sex: Female vs. Male | 0.113 | 0.133 | 0.302 |
|  | Fazekas score: CSVD+ vs. CSVD− | −0.009 | 0.900 | −0.023 |
|  | Age (years) | −0.484 | **<0.001** | −0.041 |
|  | Reynolds cardiovascular risk score | 0.080 | 0.392 | 0.014 |
| **Total Distal Length** | HIV status: Positive vs. Negative | −0.173 | **0.021** | −0.370 |
|  | Sex: Female vs. Male | 0.094 | 0.246 | 0.233 |
|  | Fazekas score: CSVD+ vs. CSVD− | 0.035 | 0.660 | −0.023 |
|  | Age (years) | −0.326 | **0.001** | −0.025 |
|  | Reynolds cardiovascular risk score | 0.024 | 0.811 | 0.004 |

**Note:** Categorical predictors are coded as binary contrasts with the reference group indicated later. For example, “HIV status: Positive vs. Negative” represents the adjusted mean difference for HIV-positive participants compared to HIV-negative participants. Standardized estimate $\beta$ expresses the effect size in standard deviation units, allowing direct comparison of the relative strength of predictors regardless of their original measurement scale.

#### 2.3 HIV effects on vessel morphometry within the CSVD group

In the second model restricted to CSVD+ subjects, we substituted the total WMH volume in place of Fazekas scores (Supplemental Table S2). HIV+ status remained significantly associated with fewer branches (*β* = *−*0.262, *p =* 0.009, *d = −*0.563) and borderline significant for shorter distal length (*β* = –0.173, *p =* 0.090, *d =* –0.361). Age also remained significant, but only for total branch number (branches *β* = *−*0.211, *p =* 0.046, *d =* –0.021; length *β* = *−*0.267, *p =* 0.256, *d =* –0.012). Total WMH volume showed only weaker negative trends that did not reach significance (branches *β* = *−*0.135, *p =* 0.186, *d =* –0.067; length *β* = *−*0.201, *p =* 0.056, *d =* –0.098).

### 3. iCafe metrics and inflammation markers

Plasma levels of sCD14, sCD163, and osteoprotegerin, ICAM and VCAM did not differ between PLWH and PWOH. Therefore, correlations among these markers and morphometry metrics were conducted using the combined cohort.

Pearson analyses showed that total branch number was significantly and negatively correlated with sCD14 (Pearson *r = −*0.161, *p =* 0.039) and sCD163 (*r = −*0.159, *p =* 0.042) levels, but not with osteoprotegerin, ICAM, or VCAM (Figure 2). Correlation analyses between total distal length and inflammatory markers revealed non-significant negative trends for sCD14, sCD163, osteoprotegerin and no trends for ICAM and VCAM (data not shown).

**Figure 2.**
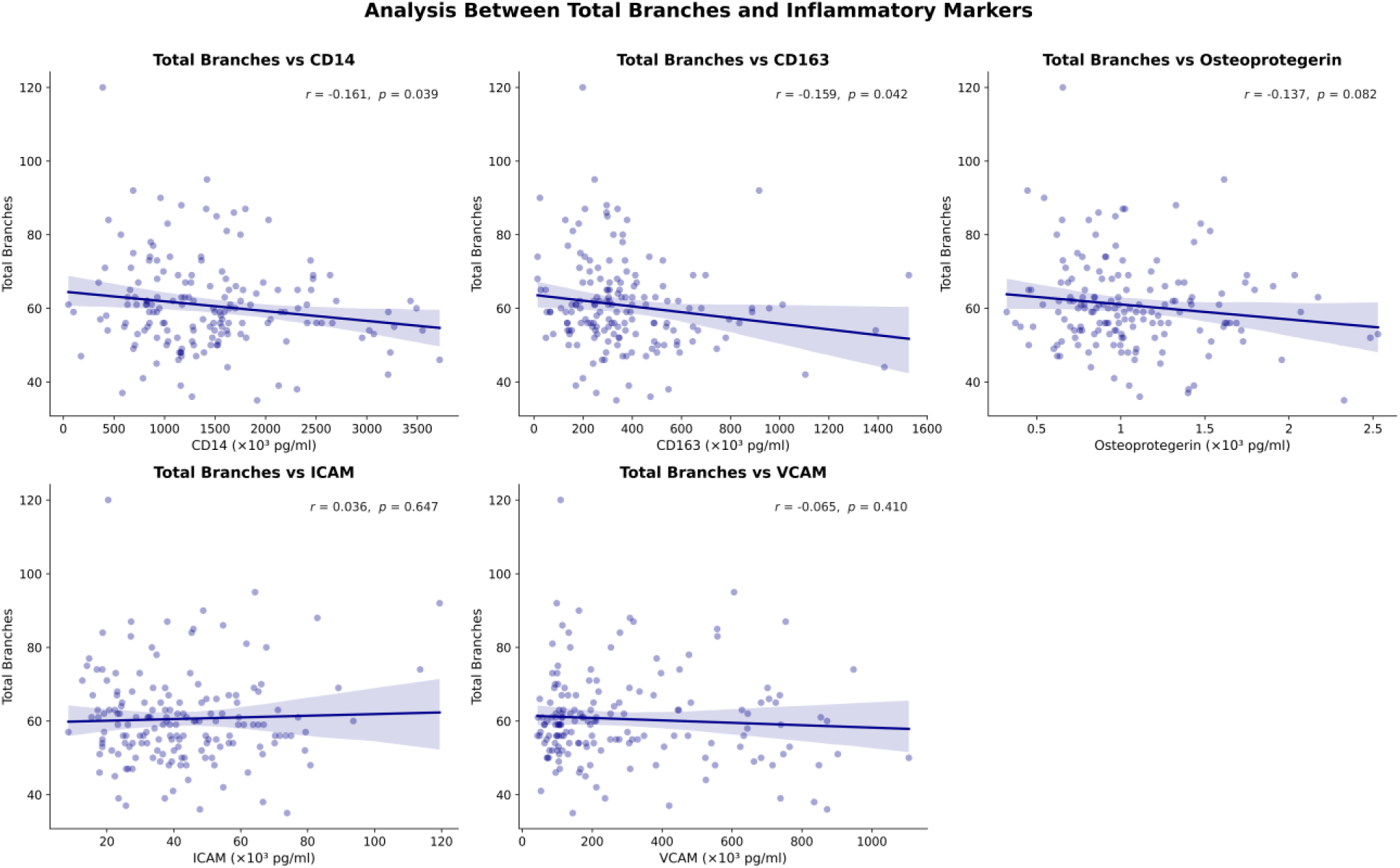
Pearson correlations between inflammatory marker levels (10^3^ pg/ml) and total branch number. Each panel corresponds to one marker: CD14, CD163, osteoprotegerin, ICAM, and VCAM. Lines indicate linear fits with 95% confidence intervals, with annotations reporting Pearson r and p-values. Total branch number was significantly and negatively correlated with sCD14 and sCD163 levels.

### 4. HIV, cerebral perfusion, and vessel morphometry

#### 4.1 Bulk CBF with respect to HIV status and cognitive function

The ANCOVA models, after adjusting for age, sex, Fazekas scores, and Reynolds risk scores, did not identify any significant associations between the global CBF and HIV status (*β* = *−*0.002, *p =* 0.984) or global CBF and total cognitive z-scores (*β* =0.035, *p =* 0.642).

However, Pearson correlation analyses between global CBF and cognition revealed significant trends with respect to PLWH, while no significant correlations between global CBF and cognitive performance were found in PWOH (Figure 3). Specifically, global CBF was positively correlated with executive function (*r =* 0.327, *p =* 0.002), processing speed (*r =* 0.297, *p =* 0.004), and attention score (*r =* 0.383, *p ≤* 0.001) domains for PLWH. In two out of three domains above, the slopes of the PLWH and PWOH cohorts were significantly different for executive function (*p =* 0.002) and processing speed (*p =* 0.008). Meanwhile, no significant correlations between CBF and total cognitive z-scores were detected (PWOH: *r = −*0.187, *p =* 0.130; PLWH: *r =* 0.135, *p =* 0.203).

**Figure 3.**
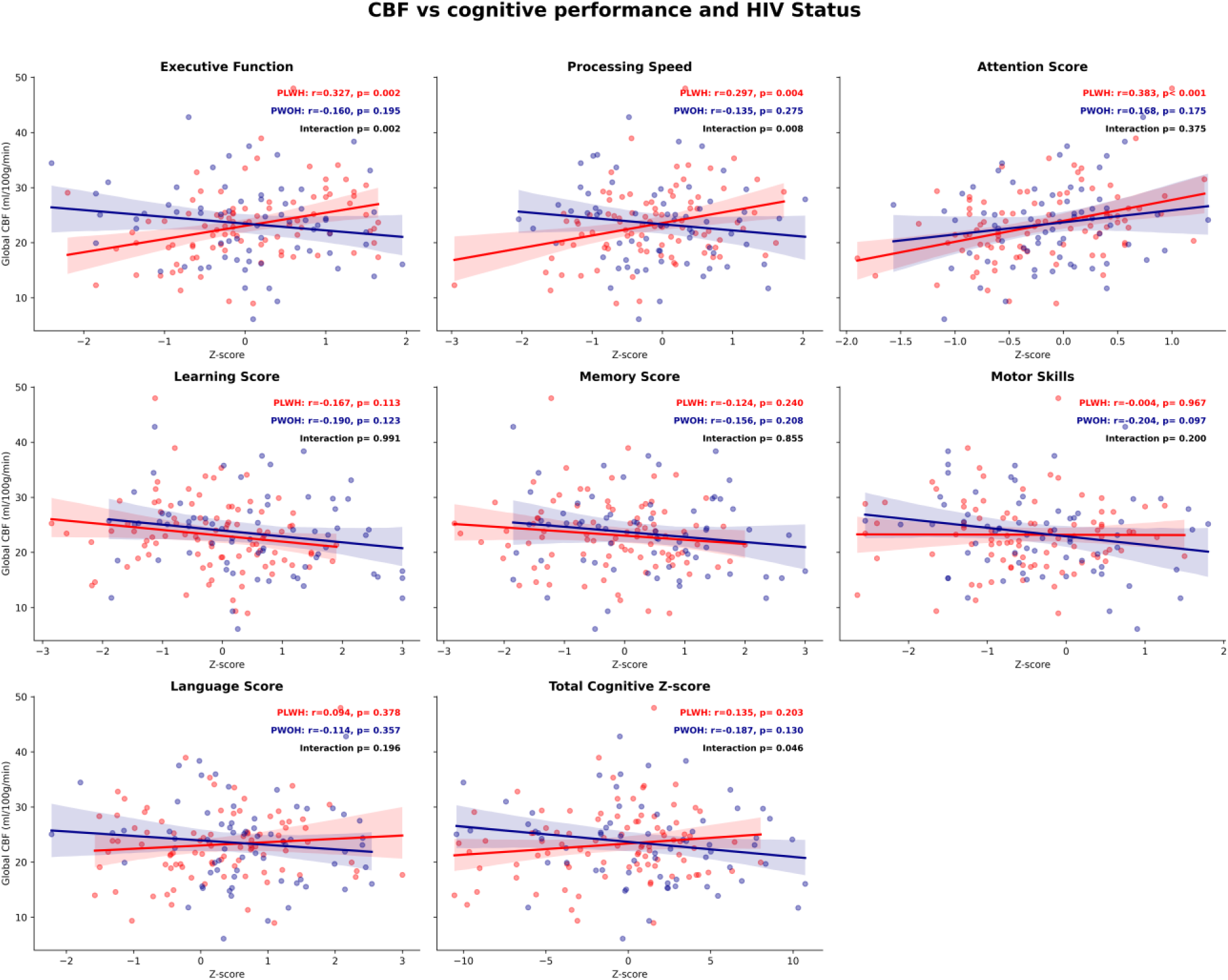
Pearson correlations between regional cerebral blood flow (CBF; ml/100g/min) and age-adjusted cognitive domains performance scores stratified by HIV status. PLWH is denoted in red, while PWOH is denoted in blue. Lines indicate linear fits with 95% confidence intervals with annotations reporting Pearson r and p-values. The p_interaction term is the p-value denoting the difference between PWOH and PLWH fitted slopes.

#### 4.2 Vascular morphometry, CBF, and age

Pearson correlation analyses between CBF and iCafe metrics revealed significant positive correlations between CBF and total number of branches for both PLWH and PWOH (0.283 ≤ *r ≤* 0.550; *p ≤* 0.05). PWOH group showed stronger associations between CBF and total branch number in SGM (*p =* 0.034) and WM (*p =* 0.050) (Supplemental Figure 1). Analysis between CBF and total distal length exhibited similar association strengths (Supplemental Figure 2). Both CBF and total branch number significantly decreased with age (*−*0.543 ≤ *r ≤ −*0.301, *p ≤* 0.004).

### 5. HIV, vessel morphometry metrics, and cognitive performance

HIV status was the strongest predictor of overall cognitive performance in both of our models, as shown in Table 3 (branch: *β* = –0.219, *p =* 0.006, *d =* –0.453; length: *β* = –0.208, *p =* 0.007, *d =* –0.435). Besides HIV, total distal length also exhibited a significant association with total cognitive scores, though with a small effect size (*β* = 0.169, *p =* 0.031, *d =* 0.001).

**Table 3:** Regression model results for branch and length metrics with total cognitive z-scores (standardized with age), adjusting for sex, HIV status, Fazekas scores, and Reynold risk scores.

| Dependent Variable | Predictor | Standardized estimate $\beta$ | p-value | Effect size d |
| --- | --- | --- | --- | --- |
| **Total Cognitive Z-score (includes Branch Number as covariate)** | HIV status: Positive vs. Negative | −0.219 | **0.006** | −0.453 |
|  | Sex: Female vs. Male | −0.082 | 0.309 | −0.197 |
|  | Fazekas score: CSVD+ vs. CSVD− | 0.037 | 0.648 | 0.085 |
|  | Total Branch Number | 0.083 | 0.303 | 0.007 |
|  | Reynolds cardiovascular risk score | −0.056 | 0.517 | −0.008 |
| **Total Cognitive Z-score (includes Total Distal Length as covariate)** | HIV status: Positive vs. Negative | −0.208 | **0.007** | −0.435 |
|  | Sex: Female vs. Male | −0.085 | 0.285 | −0.208 |
|  | Fazekas score: CSVD+ vs. CSVD− | 0.032 | 0.692 | 0.074 |
|  | Total distal length | 0.169 | **0.031** | 0.001 |
|  | Reynolds cardiovascular risk score | −0.044 | 0.601 | −0.007 |

**Note:** Analyses were performed using a regression model with either Total Branch Number (top model) or Total distal length (bottom model) entered as the primary vascular morphometric covariate, adjusting for HIV status, sex, Fazekas CSVD status, and Reynolds cardiovascular risk score. Standardized $\beta$ values are presented for comparability across predictors. Significant associations ( $p < 0.05$ ) are indicated in **bold**.

Sex, Fazekas scores, Reynolds cardiovascular risk, and total branch number did not show significant associations in the adjusted models.

At the combined cohort level, branch number and length showed significant associations with several cognitive domains, albeit with smaller effect size magnitudes. Total branch number only showed significant associations with the language domain (*β* = 0.180, *p =* 0.026, *d =* 0.015). In contrast, total distal length metrics significantly correlated with the total cognitive z-scores (*β* = 0.169, *p =* 0.031, *d =* 0.001), attention (*β* = 0.234, *p =* 0.003, *d =* 0.003), and language (*β* = 0.185, *p =* 0.019, *d =* 0.001) domains. More details are included in Supplemental Table 3.

When separated by HIV status, the Pearson analysis (Figure 4) showed significant positive trends between iCAFE distal length metrics and cognitive performance across multiple domains.

**Figure 4.**
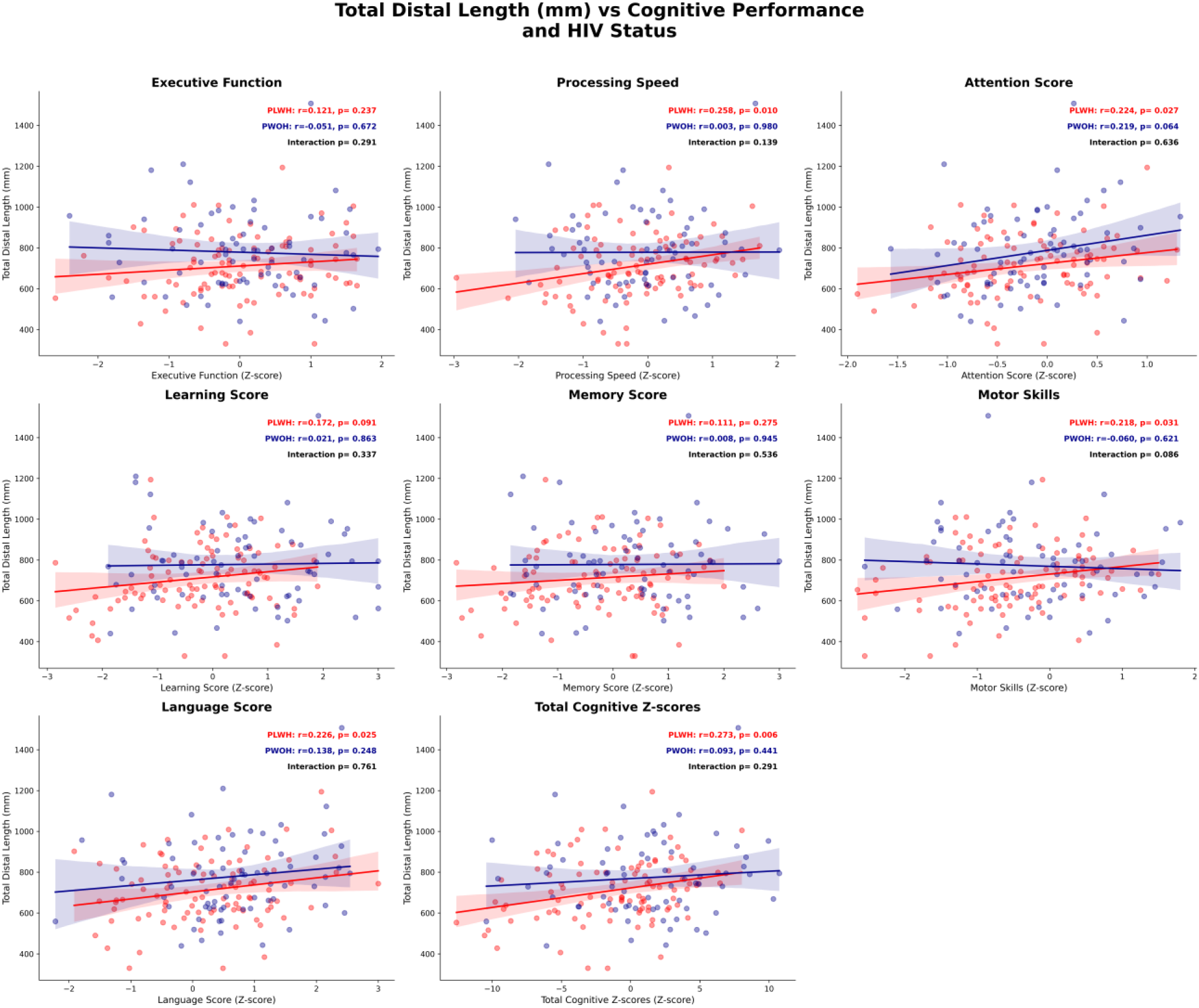
Pearson correlations between total distal length iCafe metrics in mm and age-adjusted subdomain cognitive performance scores stratified by HIV status. PLWH is denoted in red, while PWOH is denoted in blue. Lines indicate linear fits with 95% confidence intervals with annotations report Pearson r and p-values. The p_interaction term is the p-value denoted the difference between PWOH and PLWH fitted slope. Each cognitive domain is presented in a subfigure, with the total cognitive z-scores being the last panel.

Among PLWH, total distal length was correlated with processing speed (*r =* 0.258, *p =* 0.010), attention (*r =* 0.224, *p =* 0.027), motor skills (*r =* 0.218, *p =* 0.031), and language (*r =* 0.226, *p =* 0.025). Furthermore, total cognitive z-score was significantly correlated with total distal length, but only for PLWH (*r =* 0.273, *p =* 0.006). No significant correlations were observed between iCAFE total branch or total distal length metrics vs cognitive domains for PWOH.

## Discussion

The pathomechanisms of cognitive impairment in PLWH are multifactorial, especially in the aging HIV population. In this study, we examined the interplay among vessel morphological features, HIV status, and inflammatory markers, and their relationships with cognitive performance. We have shown that total branch number and total distal length were lower in the PLWH cohort compared to the PWOH group, with moderate effect sizes (branch: *β* = *−*0.224, *p =* 0.001, *d = −*0.517; length: *β* = *−*0.173, *p =* 0.021, *d = −*0.370). These vascular features were, as expected, significantly associated with age independently of HIV status (branch: *β* = *−*0.484, *p <* 0.001, *d =* –0.041; length: *β* = *−*0.326, *p =* 0.001, *d =* –0.025). Therefore, the results support the plausibility of HIV-associated contribution to vascular remodeling, independent of the effects of aging.

Systemic inflammation persists in PLWH, even with undetectable viral loads ^21^. Given the role that monocytes and endothelial cells play in vascular remodeling ^22,23^, we quantified plasma levels of monocyte products (sCD14 and sCD163) and endothelial markers (ICAM, VCAM, osteoprotegerin). From previous studies, including our own^24^, we would have expected higher plasma levels of sCD14 in PLWH compared to PWOH, but this was not the case in this relatively small cohort. However, we found significantly negative correlations between the total number of branches and sCD14 and sCD163 levels when the two groups were combined. Higher sCD14 level has been associated with higher mortality in PLWH^25^ and cardiovascular events in both PLWH and PWOH^26^ ^27^. In the general population, sCD14 has also been previously associated with cerebrovascular disease^28^ and vascular dementia^29^. Similarly, CD163 appears to be involved in cerebral small vessel disease^30^, HIV disease progression and increased cardiovascular disease mortality ^31,32^. However, the correlation strengths are smaller in magnitude with sCD14 (*r = −*0.161, *p =* 0.039) and sCD163 (*r = −*0.159, *p =* 0.042), suggesting that inflammation is only one of many factors influencing vascular remodeling.

In TOF-MRA, vascular contrast arises from the unsaturated spins within fresh blood entering the imaging volume and tracing out the vascular trees. Although branch number and total distal length captured from MRA images are not direct measurements of blood flow, the decrease in these morphometric features reflects loss of vascular integrity that may influence cerebral function^12,15,33–35^. Particularly, total branch number and vascular length were highlighted as important metrics for quantifying vascular damage in aging populations and in individuals with atherosclerosis or Alzheimer’s disease^11,12,15,36^. Consistent with these observations, we found that both total branch number and total distal length showed significant correlations with CBF in both white and gray matter regions (0.28 ≤ *r ≤* 0.55, Supplemental Figure 1).

Lower CBF is associated with cognitive impairment in patients with Alzheimer Disease (AD) and lacunar infarction^37–39^. While lower CBF level is correlated with HIV status, direct associations between CBF and cognitive performance have not been uniformly proven^40,41^. Petersen et al., (2021) noted that higher viral load is associated with both lower CBF and accelerated aging, reduced language skill, and motor function^40^. Burdo (2023) detected that higher numbers of chronic immune cell markers associated with viral infection (CD4/CD8 ratio, CD16+ monocytes) were associated with reduced brain volume^42^. CD14 marker, in particular, was associated with reduced CBF^42^. These observations suggest that chronic inflammation may affect CBF, among other contributing factors in HIV-associated cognitive impairment.

In our findings, stronger correlations between CBF and cognition were observed in PLWH (0.297 ≤ *r ≤* 0.383; *p ≤* 0.05), suggesting that alterations in perfusion may contribute to cognitive function. Of interest, among the cognitive domains affected were executive function and processing speed, two domains known to be involved with vascular cognitive impairment^43^. Nevertheless, the independent contribution of vascular morphometry to cognition, while statistically significant (*β* = 0.169, *p =* 0.031, *d =* 0.001), was much smaller in magnitude despite being highly associated with CBF. HIV status remained the strongest predictor of both vascular morphometry (*d =* –0.517) and cognitive performance (*d =* –0.435), with medium effect sizes. The relationships examined in our analyses between CBF, vessel length, and HIV support the possibility that HIV-associated neuroinflammation may contribute concomitantly to cognitive dysfunction and vascular remodeling. Furthermore, reductions in branch number and total distal length may reflect proximal changes in microvascular circulation in the distal regions of the CoW, leading to sluggish flow and loss of signal intensity. In this scenario, bulk perfusion flow measured by CBF may still appear preserved, with a reduction in cerebral flow emerging later once significant brain injury has accumulated, as in the case of AD^36,38^.

In our split-cohort analysis shown in Figure 4, higher total distal length in PLWH was associated with higher age-adjusted cognitive performance scores, as reflected in the total cognitive z-score (*r =*0.273, *p =* 0.006). Particularly, higher total distal length was correlated with higher scores in processing speed (*r =* 0.258, *p =* 0.010), attention (*r =* 0.224, *p =* 0.027), motor skills (*r =* 0.218, *p =* 0.031), as well as language (*r =* 0.226, *p =* 0.025). This is consistent with the observation of reduced distal length and cognitive decline in other neurodegenerative disorders, such as Alzheimer’s disease^36^. Yet, no such parallel was found for PWOH (Figure 4). In our marginal analysis of the combined cohort, total distal length metrics remained significantly correlated with the total cognitive z-scores, but with attenuated effect sizes (total z-score: *β* = 0.169, *p =* 0.031, *d =* 0.001; attention: *β* = 0.234, *p =* 0.003, *d =* 0.003; language: *β* = 0.185, *p =* 0.019, *d =* 0.001). The absence of correlation between vascular metrics and cognitive performance in PWOH is likely due to a ceiling effect in cognitive performance (limited range of cognitive performance).

This study has several limitations. As a semi-automatic imaging technique, iCafe results could be affected by image artifacts, low resolution, noise, and operator errors. Additionally, other measures of vascular integrity were not available, such as arterial compliance, or arterial stenosis^12^. We also did not include a hemodynamic challenge such as CO2, hyperventilation, or a task that would have further confirmed an impaired vascular regulation. Furthermore, the study demographics are skewed toward male subjects, although this imbalance is accounted for in our regression models.

Our study is cross-sectional. Thus, it does not establish a causal relationship between changes in vascular morphometry, inflammation, and cognition. In addition, the image resolution of MRA does not capture microvascular circulation, an essential component more directly linked to cognition. Moreover, the smaller effect sizes observed for vascular morphometry as a predictor of cognition suggest that a larger cohort and a better segmentation technique may be needed to precisely characterize this association. Finally, future longitudinal, multimodal studies should include higher MRA resolution and advanced MR modalities to capture blood-brain barrier and microvascular perfusion metrics (for example, diffusion prepared pseudo continuous arterial spin labeling and intravoxel incoherent motion^44,45^) to better understand the impact of perfusion on cognition within the context of HIV infection.

## Conclusion

In this study, we applied a quantitative approach to examine the changes in vascular morphometry in the context of HIV and cognitive impairment. Independent of age, sex, cardiovascular risks, and CSVD status, HIV was the strongest predictor of vascular changes as evidenced by the significantly reduced total branch number and total distal vessel length. Age was also significantly associated with vascular metrics, independent of HIV status. In the combined cohort, elevated plasma levels of monocyte activation were associated with reduced branch numbers and shorter total distal length (to a weaker extent), suggesting a possible inflammatory contribution to vascular remodeling. Among PLWH, lower total distal length was associated with lower cognitive performance (total cognitive z-scores, attention, speed, motor skills, and language) and with reduced CBF. No significant associations between iCafe metrics and cognition were found in the PWOH cohort.

Together, our results suggest that the presence of cerebrovascular dysfunction, as assessed by vascular morphometry, contributes to HIV-associated cognitive impairment. Among potential factors that can alter vascular morphometry, monocyte immune activation may play a role, though larger longitudinal studies are needed to establish directionality and significance.

## Data Availability

Manuscript data will be available upon request.

## Acknowledgments

We acknowledged our research coordinator, Allica Tyrell, for her dedication in managing the data for this research.

## Supplemental Material

- Supplemental Figure 1: Stratified correlation plots between iCAFE branch number metrics and CBF, with additional age analysis
- Supplemental Figure 2: Stratified correlation plots between iCAFE total distal length metrics and CBF, with additional age analysis
- Table S1. Group comparisons (PLWH vs. PWOH) for all vascular morphometry metrics.
- Table S2. Regression results for predictors of total branch number and total distal length within the PLWH with Fazekas scores >0.
- Table S3. Regression results for total branch and total distal length with respect to subdomain cognitive scores and total cognitive z-scores

## Non-standard Abbreviation and Acronyms

**1) Circle of Willis (CoW) abbreviations**

ACA: Anterior cerebral artery
PCA: Posterior cerebral artery
MCA: Middle cerebral artery
ICA: Internal carotid artery
BA: Basilar artery
VA: Vertebral artery
M1: Vessel in the first bifurcation of MCA
M2: Vessel in the second bifurcation of MCA
M3: Vessel in the third bifurcation of MCA
P1: Vessel in the first bifurcation of PCA
P2: Vessel in the second bifurcation of PCA
A1: Vessel in the first bifurcation of ACA
A2: Vessel in the second bifurcation of ACA
L: Anatomical left
R: Anatomical right

**2) Cortical and subcortical gray matter abbreviations:**

ACC: Anterior cingulate cortex
AMY: Amygdala
CAU: Caudate nucleus
HIP: Hippocampus
PAL: Globus Pallidus
PUT: Putamen
THA: Thalamus
SGM: Subcortical gray matter
CGM: Cortical gray matter
GWM: Generalized white matter

## REFERENCES

1. Alford K, Vera JH. Cognitive Impairment in people living with HIV in the ART era: A Review. British Medical Bulletin. 2018;127:55–68. doi: 10.1093/bmb/ldy019

2. Bandera A, Taramasso L, Bozzi G, Muscatello A, Robinson JA, Burdo TH, Gori A. HIV-Associated Neurocognitive Impairment in the Modern ART Era: Are We Close to Discovering Reliable Biomarkers in the Setting of Virological Suppression? Frontiers in Aging Neuroscience. 2019;11:187. doi: 10.3389/fnagi.2019.00187

3. Nasi M, Biasi SD, Gibellini L, Bianchini E, Pecorini S, Bacca V, Guaraldi G, Mussini C, Pinti M, Cossarizza A. Ageing and inflammation in patients with HIV infection. Clinical and Experimental Immunology. 2016 Aug 9;187. doi: 10.1111/cei.12814

4. D’Abramo A, Zingaropoli MA, Oliva A, D’Agostino C, Al Moghazi S, De Luca G, Iannetta M, Mastroianni CM, Vullo V. Immune Activation, Immunosenescence, and Osteoprotegerin as Markers of Endothelial Dysfunction in Subclinical HIV-Associated Atherosclerosis. Mediators of Inflammation. 2014;2014:192594. doi: 10.1155/2014/192594

5. Gutierrez J, Albuquerque ALA, Falzon L. HIV infection as vascular risk: A systematic review of the literature and meta-analysis. PLOS ONE. 2017;12:e0176686. doi: 10.1371/journal.pone.0176686

6. Gutierrez J, Elkind MSV, Petito C, Chung DY, Dwork AJ, Marshall RS. The contribution of HIV infection to intracranial arterial remodeling: a pilot study. Neuropathology. 2013;33:256–263. doi: 10.1111/j.1440-1789.2012.01358.x

7. Kearns A, Gordon J, Burdo TH, Qin X. HIV-1-Associated Atherosclerosis: Unraveling the Missing Link. Journal of the American College of Cardiology. 2017;69:3084–3098. doi: 10.1016/j.jacc.2017.05.012

8. Moulignier A, Savatovsky J, Assoumou L, Lescure F-X, Lamirel C, Godin O, Valin N, Tubiana R, Canestri A, Roux P, et al. Silent Cerebral Small-Vessel Disease Is Twice as Prevalent in Middle-Aged Individuals With Well-Controlled, Combination Antiretroviral Therapy–Treated Human Immunodeficiency Virus (HIV) Than in HIV-Uninfected Individuals. Clinical Infectious Diseases. 2018;66:1762–1769. doi: 10.1093/cid/cix1075

9. Edelman RR, Koktzoglou I. Non-Contrast MR Angiography:An Update. Journal of magnetic resonance imaging : JMRI. 2019/02;49. doi: 10.1002/jmri.26288

10. Chen L, Mossa-Basha M, Balu N, Canton G, Sun J, Pimentel K, Hatsukami TS, Hwang JN, Yuan C. Development of a quantitative intracranial vascular features extraction tool on 3D MRA using semiautomated open-curve active contour vessel tracing. Magn Reson Med. 2018;79:3229–3238. doi: 10.1002/mrm.26961

11. Chen L, Sun J, Hippe DS, Balu N, Yuan Q, Yuan I, Zhao X, Li R, He L, Hatsukami TS, et al. Quantitative Assessment of the Intracranial Vasculature in an Older Adult Population using iCafe (intraCranial Artery Feature Extraction). Neurobiology of Aging. 2019;79:59–65. doi: 10.1016/j.neurobiolaging.2019.02.027

12. Chen Z, Gould A, Geleri DB, Balu N, Chen L, Chu B, Pimentel K, Canton G, Hatsukami TS, Yuan C. Associations of intracranial artery length and branch number on non-contrast enhanced MRA with cognitive impairment in individuals with carotid atherosclerosis. Sci Rep. 2022;12:7456. doi: 10.1038/s41598-022-11418-y

13. Cheng H, Teng J, Jia L, Xu L, Yang F, Li H, Ling C, Liu W, Li J, Li Y, et al. Association between morphologic features of intracranial distal arteries and brain atrophy indexes in cerebral small vessel disease: a voxel-based morphometry study. Front Neurol. 2023;14:1198402. doi: 10.3389/fneur.2023.1198402

14. Gould A, Chen Z, Geleri DB, Balu N, Zhou Z, Chen L, Chu B, Pimentel K, Canton G, Hatsukami T, et al. Vessel length on SNAP MRA and TOF MRA is a potential imaging biomarker for brain blood flow. Magnetic Resonance Imaging. 2021;79:20–27. doi: 10.1016/j.mri.2021.02.012

15. Deshpande A, Elliott J, Kari N, Jiang B, Michel P, Toosizadeh N, Fahadan PT, Kidwell C, Wintermark M, Laksari K. Novel imaging markers for altered cerebrovascular morphology in aging, stroke, and Alzheimer’s disease. Journal of Neuroimaging. 2022;32:956–967. doi: 10.1111/jon.13023

16. Murray K, Uddin M, Tivarus M, Sahin B, Wang H, Singh M, Qiu X, Wang L, Spincemaille P, Wang Y, et al. Increased Risk for Cerebral Small Vessel Disease is Associated with Quantitative Susceptibility Mapping in HIV Infected and Uninfected Individuals. NeuroImage: Clinical. 2021;32:102786. doi: 10.1016/j.nicl.2021.102786

17. Chen L, Mossa-Basha M, Balu N, Canton G, Sun J, Pimentel K, Hatsukami TS, Hwang J-N, Yuan C. Development of a quantitative intracranial vascular features extraction tool on 3D MRA using semiautomated open-curve active contour vessel tracing. Magnetic Resonance in Medicine. 2018;79:3229–3238. doi: 10.1002/mrm.26961

18. M J, CF B, TE B, MW W, SM S. FSL - PubMed. NeuroImage. 08/15/2012;62. doi: 10.1016/j.neuroimage.2011.09.015

19. Fazekas F, Chawluk JB, Alavi A, Hurtig HI, Zimmerman RA. MR signal abnormalities at 1.5 T in Alzheimer’s dementia and normal aging. AJR American journal of roentgenology. 1987;149:351–356. doi: 10.2214/ajr.149.2.351

20. Manjón JV, Coupé P. Frontiers | volBrain: An Online MRI Brain Volumetry System. Frontiers in Neuroinformatics. 2016/07/27;10. doi: 10.3389/fninf.2016.00030

21. Masyuko SJ, Page ST, Polyak SJ, Kinuthia J, Osoti AO, Otieno FC, Kibachio JM, Mogaka JN, Macharia PM, Chohan BH, et al. Human Immunodeficiency Virus Is Associated With Higher Levels of Systemic Inflammation Among Kenyan Adults Despite Viral Suppression. Clinical Infectious Diseases: An Official Publication of the Infectious Diseases Society of America. 2020;73:e2034–e2042. doi: 10.1093/cid/ciaa1650

22. Pereyra F, Lo J, Triant VA, Wei J, Buzon MJ, Fitch KV, Hwang J, Campbell JH, Burdo TH, Williams KC, et al. Increased Coronary Atherosclerosis and Immune Activation in HIV-1 Elite Controllers. AIDS. 2012;26:2409–2412. doi: 10.1097/QAD.0b013e32835a9950

23. Kim K-W, Ivanov S, Williams JW. Monocyte Recruitment, Specification, and Function in Atherosclerosis. Cells. 2020;10:15. doi: 10.3390/cells10010015

24. Singh MV, Uddin MN, Covacevich Vidalle M, Sutton KR, Boodoo ZD, Peterson AN, Tyrell A, Tivarus ME, Wang HZ, Sahin B, et al. Non-classical monocyte levels correlate negatively with HIV-associated cerebral small vessel disease and cognitive performance. Frontiers in cellular and infection microbiology. 2024;14:1405431. doi: 10.3389/fcimb.2024.1405431

25. Sandler NG, Wand H, Roque A, Law M, Nason MC, Nixon DE, Pedersen C, Ruxrungtham K, Lewin SR, Emery S, et al. Plasma Levels of Soluble CD14 Independently Predict Mortality in HIV Infection. J Infect Dis. 2011;203:780–790. doi: 10.1093/infdis/jiq118

26. Ntsekhe M, Baker JV. Cardiovascular Disease Among Persons Living With HIV: New Insights Into Pathogenesis and Clinical Manifestations in a Global Context. Circulation. 2023;147:83–100. doi: 10.1161/CIRCULATIONAHA.122.057443

27. Al-Kindi SG, Buzkova P, Shitole SG, Reiner AP, Garg PK, Gottdiener JS, Psaty BM, Kizer JR. Soluble CD14 and Risk of Heart Failure and its Subtypes in Older Adults. J Card Fail. 2020;26:410–419. doi: 10.1016/j.cardfail.2020.03.003

28. Olson NC, Koh I, Reiner AP, Judd SE, Irvin MR, Howard G, Zakai NA, Cushman M. Soluble CD14, Ischemic Stroke, and Coronary Heart Disease Risk in a Prospective Study: The REGARDS Cohort. Journal of the American Heart Association. 2020;9:e014241. doi: doi:10.1161/JAHA.119.014241

29. Pase MP, Himali JJ, Beiser AS, DeCarli C, McGrath ER, Satizabal CL, Aparicio HJ, Adams HHH, Reiner AP, Longstreth WT, et al. Association of CD14 with incident dementia and markers of brain aging and injury. Neurology. 2020;94:e254–e266. doi: doi:10.1212/WNL.0000000000008682

30. Chen Y, Lu P, Wu S, Yang J, Liu W, Zhang Z, Xu Q. CD163-Mediated Small-Vessel Injury in Alzheimer’s Disease: An Exploration from Neuroimaging to Transcriptomics. Int J Mol Sci. 2024;25. doi: 10.3390/ijms25042293

31. Burdo TH, Lo J, Abbara S, Wei J, DeLelys ME, Preffer F, Rosenberg ES, Williams KC, Grinspoon S. Soluble CD163, a Novel Marker of Activated Macrophages, Is Elevated and Associated With Noncalcified Coronary Plaque in HIV-Infected Patients. J Infect Dis. 2011;204:1227–1236. doi: 10.1093/infdis/jir520

32. Durda P, Raffield LM, Lange EM, Olson NC, Jenny NS, Cushman M, Deichgraeber P, Grarup N, Jonsson A, Hansen T, et al. Circulating Soluble CD163, Associations With Cardiovascular Outcomes and Mortality, and Identification of Genetic Variants in Older Individuals: The Cardiovascular Health Study. J Am Heart Assoc. 2022;11:e024374. doi: 10.1161/JAHA.121.024374

33. Chen L, Sun J, Hippe DS, Balu N, Yuan Q, Yuan I, Zhao X, Li R, He L, Hatsukami TS, et al. Quantitative assessment of the intracranial vasculature in an older adult population using iCafe. Neurobiology of Aging. 2019;79:59–65. doi: 10.1016/j.neurobiolaging.2019.02.027

34. Chen Z, Chen L, Shirakawa M, Liu W, Ortega D, Chen J, Balu N, Trouard T, Hatsukami TS, Zhou W, et al. Intracranial vascular feature changes in time of flight MR angiography in patients undergoing carotid revascularization surgery. Magnetic Resonance Imaging. 2021;75:45–50. doi: 10.1016/j.mri.2020.10.004

35. Chen Z, Liu W, Balu N, Chen L, Ortega D, Huang X, Hatsukami TS, Yang J, Yuan C. Associations of Intracranial Artery Length and Branch Number on Time-of-Flight MRA With Cognitive Impairment in Hypertensive Older Males. J Magn Reson Imaging. 2024. doi: 10.1002/jmri.29242

36. Kim T, Zhang K, Zhu T, Song I-U, Na S, Hippe DS, Canton G, Hatsukami T, Yuan C, Mossa-Basha M, et al. Quantitative analysis of cerebral vasculature and its clinical effect on cognitive change in Alzheimer’s dementia and mild cognitive impairment. Journal of the Neurological Sciences. 2025;476:123598. doi: 10.1016/j.jns.2025.123598

37. Kitagawa K, Oku N, Kimura Y, Yagita Y, Sakaguchi M, Hatazawa J, Sakoda S. Relationship between cerebral blood flow and later cognitive decline in hypertensive patients with cerebral small vessel disease. Hypertens Res. 2009;32:816–820. doi: 10.1038/hr.2009.100

38. Leeuwis AE, Benedictus MR, Kuijer JPA, Binnewijzend MAA, Hooghiemstra AM, Verfaillie SCJ, Koene T, Scheltens P, Barkhof F, Prins ND, et al. Lower cerebral blood flow is associated with impairment in multiple cognitive domains in Alzheimer’s disease. Alzheimers Dement. 2017;13:531–540. doi: 10.1016/j.jalz.2016.08.013

39. Leeuwis AE, Smith LA, Melbourne A, Hughes AD, Richards M, Prins ND, Sokolska M, Atkinson D, Tillin T, Jäger HR, et al. Cerebral Blood Flow and Cognitive Functioning in a Community-Based, Multi-Ethnic Cohort: The SABRE Study. Frontiers in Aging Neuroscience. 2018;10. doi: 10.3389/fnagi.2018.00279

40. Petersen KJ, Metcalf N, Cooley S, Tomov D, Vaida F, Paul R, Ances BM. Accelerated Brain Aging and Cerebral Blood Flow Reduction in Persons With Human Immunodeficiency Virus. Clinical Infectious Diseases: An Official Publication of the Infectious Diseases Society of America. 2021;73:1813–1821. doi: 10.1093/cid/ciab169

41. Su T, Mutsaerts HJMM, Caan MWA, Wit FWNM, Schouten J, Geurtsen GJ, Sharp DJ, Prins M, Richard E, Portegies P, et al. Cerebral blood flow and cognitive function in HIV-infected men with sustained suppressed viremia on combination antiretroviral therapy. AIDS. 2017;31:847. doi: 10.1097/QAD.0000000000001414

42. Burdo TH, Robinson JA, Cooley S, Smith MD, Flynn J, Petersen KJ, Nelson B, Westerhaus E, Wisch J, Ances BM. Increased Peripheral Inflammation Is Associated With Structural Brain Changes and Reduced Blood Flow in People With Virologically Controlled HIV. J Infect Dis. 2023;228:1071–1079. doi: 10.1093/infdis/jiad229

43. Hamilton OKL, Backhouse EV, Janssen E, Jochems ACC, Maher C, Ritakari TE, Stevenson AJ, Xia L, Deary IJ, Wardlaw JM. Cognitive impairment in sporadic cerebral small vessel disease: A systematic review and meta-analysis. Alzheimer’s & Dementia. 2021;17:665–685. doi: 10.1002/alz.12221

44. Kerkhofs D, Wong SM, Zhang E, Staals J, Jansen JFA, van Oostenbrugge RJ, Backes WH. Baseline Blood-Brain Barrier Leakage and Longitudinal Microstructural Tissue Damage in the Periphery of White Matter Hyperintensities. Neurology. 2021;96:e2192–e2200. doi: 10.1212/WNL.0000000000011783

45. Paschoal AM, Leoni RF, dos Santos AC, Paiva FF. Intravoxel incoherent motion MRI in neurological and cerebrovascular diseases. NeuroImage : Clinical. 2018;20:705–714. doi: 10.1016/j.nicl.2018.08.030

