## Supplemental Figures 1 and 2 for "Morphological feature remodeling of intracranial arteries in the context of inflammation and HIV-associated cognitive impairment"

### SUPPLENTAL FIGURES

- Supplemental Figure 1: Stratified correlation plots between iCAFE branch number metrics and CBF, with additional age analysis
- Supplemental Figure 2: Stratified correlation plots between iCAFE total distal length metrics and CBF, with additional age analysis

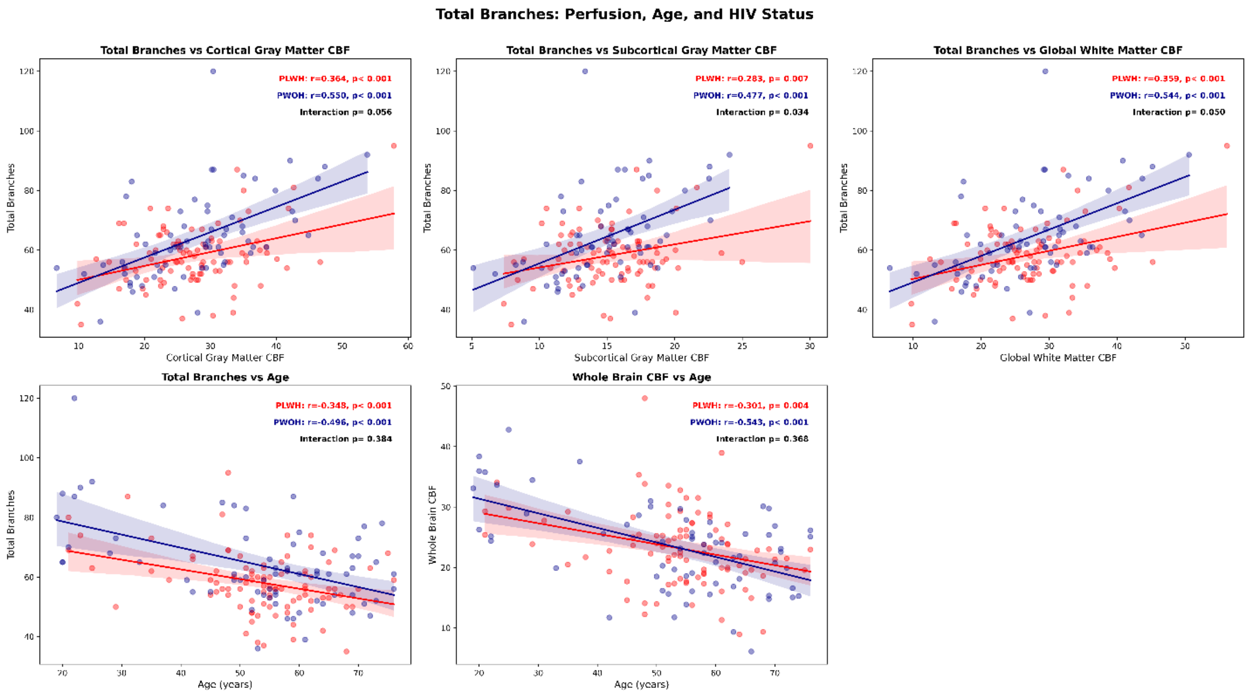


***Supplementary Figure 1****. Pearson correlations between regional cerebral blood flow (CBF; ml/100g/min), iCafe branch number, and age stratified by HIV status. PLWH is denoted in red, while PWOH is denoted in blue. The top row displays the trends between CBF and the iCafe total branch number. The bottom row displays the relationship between total branch number and CBF with respect to age. Lines indicate linear fits with 95% confidence intervals with annotations report Pearson r and p-values. The p_interaction term is the p-value denoted the difference between PWOH and PLWH fitted slope. Avg: Average; GWM: Global white matter; CGM: Cortical gray matter; SGM: subcortical gray matter.*

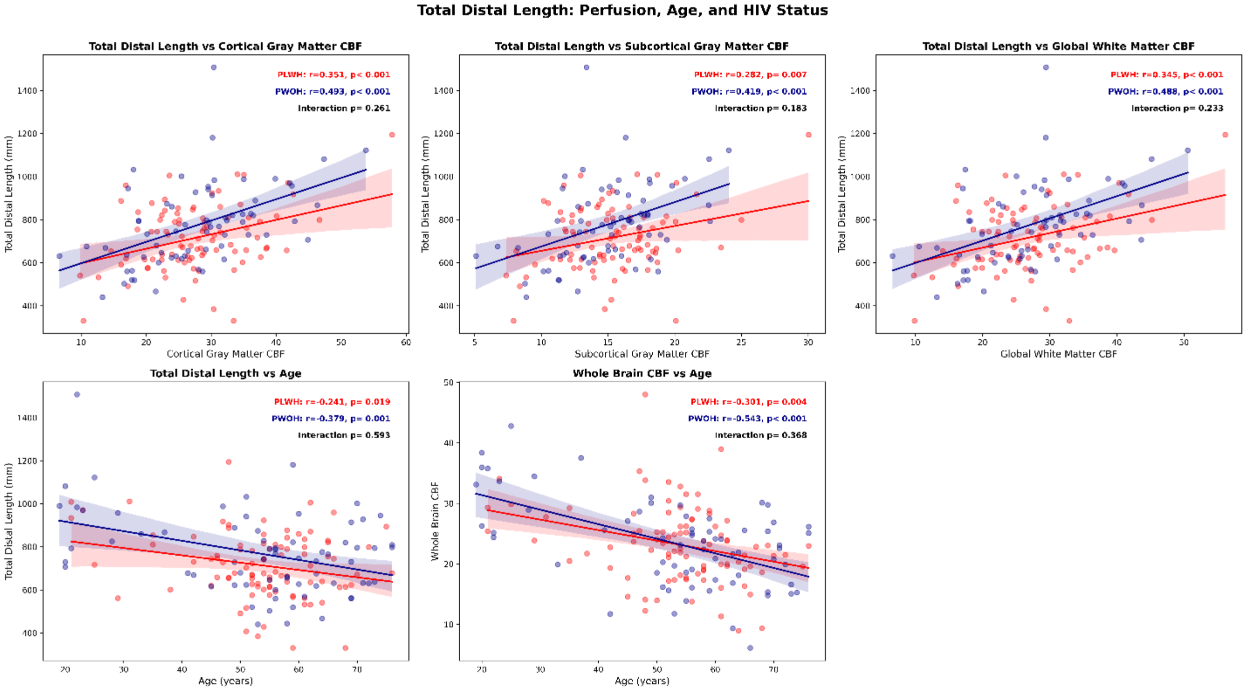


***Supplementary Figure 2****. Pearson correlations between regional cerebral blood flow (CBF; ml/100g/min), iCafe total distal length (mm), and age stratified by HIV status. PLWH is denoted in red, while PWOH is denoted in blue. Top row: the trends between CBF and the iCafe metrics total distal length. The bottom row: the relationship between iCAFE total distal length and CBF with respect to age. Lines indicate linear fits with 95% confidence intervals with annotations report Pearson r and p-values. The p_interaction term is the p-value denoted the difference between PWOH and PLWH fitted slope. Avg: Average; GWM: Global white matter; CGM: Cortical gray matter; SGM: subcortical gray matter.*
