## Supplemental Tables 1-3 for "Morphological feature remodeling of intracranial arteries in the context of inflammation and HIV-associated cognitive impairment"

**SUPPLENTAL TABLES**

**List of Supplemental Tables**

- Table S1. Group comparisons (PLWH vs. PWOH) for all vascular morphometry metrics.
- Table S2. Regression results for predictors of total branch number and total distal length within the PLWH with Fazekas scores >0.
- Table S3. Regression results for total branch and total distal length with respect to subdomain cognitive scores and total cognitive z-scores

**Table S1.** Group comparisons (PLWH vs. PWOH) for all vascular morphometry metrics. Negative mean differences indicate lower values in PLWH. Welch’s t-test is used for unadjusted analysis. Other metrics are not included due to the lack of significant findings.

| **Feature** | **Mean + SD (PLWH)** | **Mean + SD (PWOH)** | **t-value** | **p-value** | **Effect**  **Size d** |
| --- | --- | --- | --- | --- | --- |
| *Total branch number* | 58.41 ± 10.06 | 64.23 ± 14.26 | –2.98 | **0.003** | **–0.484** |
| Left MCA | 16.60 ± 4.28 | 18.00 ± 4.66 | –2.02 | **0.045** | **–0.316** |
| Right MCA | 15.53 ± 3.72 | 17.70 ± 4.82 | –3.21 | **0.002** | **–0.515** |
| Total MCA | 32.12 ± 6.72 | 35.70 ± 8.66 | –2.94 | **0.004** | –0.471 |
| Left PCA | 5.07 ± 2.15 | 5.75 ± 2.08 | –2.10 | **0.038** | –**0.322** |
| Right PCA | 5.23 ± 1.99 | 5.70 ± 2.43 | –1.34 | 0.182 | –0.213 |
| Total PCA | 10.30 ± 3.79 | 11.45 ± 3.83 | –1.95 | 0.053 | –0.302 |
| Left ACA | 5.09 ± 2.45 | 5.30 ± 2.69 | –0.53 | 0.599 | –0.082 |
| Right ACA | 4.78 ± 2.28 | 5.45 ± 2.67 | –1.74 | 0.084 | –0.274 |
| Total ACA | 9.87 ± 3.59 | 10.75 ± 4.35 | –1.42 | 0.159 | –0.225 |
| *Total distal vessel length (mm)* | 1426.68 ± 303.62 | 1555.56 ± 384.90 | –2.37 | **0.019** | **–0.379** |
| Left MCA+ PCA+ ACA | 767.02 ± 171.62 | 826.89 ± 186.08 | –2.14 | **0.034** | **–0.337** |
| Right MCA+ PCA+ ACA | 752.72 ± 148.79 | 822.76 ± 213.31 | –2.34 | **0.021** | **–0.391** |
| Left MCA | 496.13 ± 122.00 | 537.05 ± 121.82 | –2.18 | **0.031** | **–0.336** |
| Right MCA | 483.98 ± 107.61 | 532.36 ± 136.71 | –2.50 | **0.013** | **–0.400** |
| Left PCA | 144.28 ± 47.90 | 159.14 ± 51.63 | –1.92 | 0.056 | **–0.300** |
| Right PCA | 147.61 ± 50.59 | 160.27 ± 57.54 | –1.50 | 0.136 | **–0.236** |
| Left ACA | 126.60 ± 53.32 | 130.70 ± 56.43 | –0.48 | 0.630 | **–0.075** |
| Right ACA | 121.13 ± 50.60 | 130.13 ± 61.29 | –1.02 | 0.308 | **–0.163** |
| Left ICA | 88.79 ± 10.71 | 92.65 ± 16.19 | –1.77 | 0.079 | **–0.214** |
| Right ICA | 88.19 ± 11.37 | 91.12 ± 16.26 | –1.32 | 0.190 | **–0.290** |

***Note:*** *MCA:* *middle cerebral artery; PCA: posterior cerebral artery; ACA: anterior cerebral artery; ICA: internal carotid artery vessel length.*

***Table S2.*** *Regression results for predictors of total branch number and total distal length within the CSVD+ (Fazekas scores >0).*

| **Dependent Variable** | **Predictor** | **Standardized Estimate (β)** | **p-value** | **Effect**  **Size d** |
| --- | --- | --- | --- | --- |
| Total Branch Number | Sex: Female vs. Male | 0.109 | 0.307 | 0.248 |
|  | HIV status: Positive vs. Negative | –0.262 | **0.009** | –0.563 |
|  | Age (years) | –0.211 | **0.046** | –0.021 |
|  | Reynolds cardiovascular risk score | 0.062 | 0.619 | 0.009 |
|  | Total lesion volume (cm³) | –0.135 | 0.186 | –0.067 |
| Total distal length | Sex: Female vs. Male | 0.091 | 0.400 | 0.204 |
|  | HIV status: Positive vs. Negative | –0.173 | **0.090** | –0.361 |
|  | Age (years) | –0.267 | **0.256** | –0.012 |
|  | Reynolds cardiovascular risk score | 0.020 | 0.872 | 0.003 |
|  | Total lesion volume (cm³) | –0.201 | 0.056 | –0.098 |

***Note:*** *Categorical predictors are coded as binary contrasts with the reference group indicated later. For example, “HIV status: Positive vs. Negative” represents the adjusted mean difference for HIV-positive participants compared to HIV-negative participants. Significant p-values (< 0.05) are in* ***bold****.*

**Table S3.** *Regression results for total branch and total distal length with respect to subdomain cognitive scores and total cognitive z-scores*

| **Cognitive Domain (Z-score)** | **Vascular Metric** | **Standard**  **Estimate (β)** | **p-value** | **Effect size d** |
| --- | --- | --- | --- | --- |
| ATT | Total Branch Number | 0.142 | 0.086 | 0.012 |
|  | Total Distal Length | 0.234 | **0.003** | **0.003** |
| EXE | Total Branch Number | –0.048 | 0.562 | –0.003 |
|  | Total Distal Length | 0.032 | 0.690 | <0.001 |
| LAN | Total Branch Number | 0.180 | **0.026** | **0.015** |
|  | Total Distal Length | 0.185 | **0.019** | **0.001** |
| LEA | Total Branch Number | 0.057 | 0.470 | 0.004 |
|  | Total Distal Length | 0.115 | 0.135 | <0.001 |
| MEM | Total Branch Number | 0.052 | 0.516 | 0.004 |
|  | Total Distal Length | 0.086 | 0.267 | <0.001 |
| MOT | Total Branch Number | –0.021 | 0.799 | –0.001 |
|  | Total Distal Length | 0.067 | 0.407 | <0.001 |
| SPE | Total Branch Number | 0.058 | 0.480 | 0.005 |
|  | Total Distal Length | 0.146 | 0.068 | <0.001 |
| Total Z-Scores | Total Branch Number | 0.083 | 0.303 | 0.006 |
|  | Total Distal Length | 0.169 | **0.031** | **0.001** |

***Note:*** Values represent standardized β coefficients from separate regression models, with each cognitive domain Z-score entered as the dependent variable and either ***Total Branch Number*** or ***Total distal length*** as the primary vascular morphometric covariate. Models were adjusted for HIV status, sex, Fazekas CSVD status, and Reynolds cardiovascular risk score. Significant associations (p < 0.05) are shown in ***bold***.
